# Rate Estimation and Identification of COVID-19 Infections: Towards Rational Policy Making During Early and Late Stages of Epidemics

**DOI:** 10.1101/2020.05.22.20110585

**Authors:** Richard Beigel, Simon Kasif

**Affiliations:** Department of Computer and Information Sciences, Temple University, Philadelphia 19122; Department of Biomedical Engineering, Boston University, Boston 02215

## Abstract

Pandemics have a profound impact on our world, causing loss of life, affecting our culture and historically shaping our genetics. The response to a pandemic requires both resilience and imagination. It has been clearly documented that obtaining an accurate estimate and trends of the actual infection rate and mortality risk are very important for policy makers and medical professionals. One cannot estimate mortality rates without an accurate assessment of the number of infected individuals in the population. This need is also aligned with identifying the infected individuals so they can be properly treated, monitored and tracked. However, accurate estimation of the infection rate, locally, geographically and nationally is important independently. These infection rate estimates can guide policy makers at both state, national or world level to achieve a better management of risk to society. The decisions facing policy makers are very different during early stages of an emerging epidemic where the infection rate is low, middle stages where the rate is rapidly climbing, and later stages where the epidemic curve has flattened to a low and relatively sustainable rate. In this paper we provide relatively efficient pooling methods to both estimate infection rates and identify infected individuals for populations with low infection rates. These estimates may provide significant cost reductions for testing in rural communities, third world countries and other situations where the cost of testing is expensive or testing is not widely available. As we prepare for the second wave of the pandemic this line of work may provide new solutions for both the biomedical community and policy makers at all levels.

## Introduction

COVID-19 is a deadly disease caused by the Sars-CoV-2 RNA virus. This novel coronavirus created an epidemic of global proportions killing over 200,000 people worldwide (as of April 28^th^, 2020) and infecting millions. It also caused a medical crisis and unprecedented disruptions that long-term are likely to increase the risk for multiple-socio-economic downturns that are associated with both mortality and chronic disease.

The pandemic created many urgent problems such as the development of antiviral medications, vaccines, and inexpensive and widely available testing capability, expanding ER and ICU capabilities and much more. However, proper response to the pandemic requires estimates of the rate of infection via testing. The challenge of rapid testing has created an outstanding community response from industry and academic centers producing tests to diagnose and identify infected patients. The majority of these tests rely on PCR based techniques that are very well established. Newer tests are based on isothermal amplification and most recently CRISPR based methods using CAS-13. These tests can deliver a result in minutes to hours. High throughput sequencing is also an option for large scale testing.

In addition to the biomedical crisis, the virus has also created major challenges for policy makers that need to make life and death decisions based on ethical, clinical and economic factors of unprecedented importance. A full shutdown of the economy and forced social distancing create a colossal stress on the economy, unemployment, and collapse of many business sectors. Opening the economy would increase risk for the aging population and people with pre-existing conditions such as diabetes, cancer, cardiovascular disease, respiratory disease, and immunodeficiency.

The decisions facing policy makers are very different at the beginning of an emerging epidemic where the infection rate is low vs middle stages (when the rates are rapidly climbing) and at the later stages where the epidemic curve flattened to a low and relatively sustainable rate. During the early stages it is relatively inefficient to test millions of patients who might be suffering from symptoms caused by RSV or influenza. At the same time is it important to monitor any unexpected turns in the progression. In many rural communities the rates usually remain low except local bursts that need to be contained. Similarly in third world countries it is economically and practically impossible to perform many tests early on.

Mathematically, the problems of identifying infected individuals (**identification**) and estimating the total number of infected individuals in a given population (**infection rate**) are related but in fact can be addressed by subtly different algorithms to reduce the number of tests needed and thereby the total cost of doing testing. These methods generally rely on a well-studied area called combinatorial group testing. However, as we will demonstrate in this brief communication, estimating the number of infected individuals can be solved by novel adaptation of methods developed in theoretical computer science aimed at approximate counting. Here we refer to these methods as ACA (approximate counting algorithms). More specifically, we describe comparatively efficient methods enabling estimation of the total number of infected individuals. Intuitively, we pool samples from multiple individuals and repeatedly test these pools for the virus with a single test (or perhaps a small constant number of tests to achieve better sensitivity). We provide a detailed analysis of the accuracy of the approximate counting procedure with both theoretical analysis and simulation. As a simple example, if the infection rate in a population of 1,000,000 is around 1% we can produce an unbiased estimate of the infection rate with variance approximately 7·10^−5^ by making 20 group tests. In contrast, testing a sample of 20 individuals to estimate the infection rate would produce an estimate whose variance is 4.95·10^−4^. Our variance is approximately 7 times smaller.

In addition to rate estimation we provide a review and analysis of several identification algorithms that can be deployed in communities with low infection rates that achieve reasonable improvement over the standard algorithms for group testing that have been previously explored.

## Methods

### Section 1: Assumptions, Terminology and Rate Estimation without Identification

We first abstract the key computational problems as follows:

- Estimate the rate of the infection in the population or approximately count how many people test positive in a population of a given size with as few partially pooled tests as possible.
- Identify the people who are positive with as few partially pooled tests as possible.

We focus on these problems in the context of the population that are actively suffering from the disease, rather than post-infection and recovery. We assume the testing is done using established genomic testing procedures. While similar methods are feasible to implement to detect individuals that have or already had the disease (e.g. via antibody presence testing) we expect this number to be significantly higher and our methods are comparatively more effective for lower fractions of individuals testing positive.

**Batch Testing Assumption:** Given a set of samples from a set S of people (infected or not), it is technically feasible to form a single batch consisting of all samples and test that batch with a single test. The batch will test positive if and only if at least one person in S is positive. This assumption is reasonable for small batch sizes (e.g. 100 ≤ |S| ≤ 1000). We will provide methods to alleviate this technical restriction on batch size in the discussion.

Henceforth, n will refer to the (known) number of people (total size of the population) and k will stand for the (possibly unknown) number of people who will test positive. Thus k/N is the infection rate. We study two problems: **identification** of infected individuals and estimating k by **approximate counting**. The formulas used throughout the methods section are provided in the Supplement for convenience.

Typically, one estimates the infection rate by sampling individuals. This estimate of the rate using the sample mean, is known to be highly inaccurate when the probability of infection p is small, because the variance of the estimator is much larger than p^2^. Using group testing, we will produce estimates whose variance is asymptotically proportional to p^2^. Therefore our proposed methods are superior to sampling individuals when p is small.

We will define a random variable Y such that

- Y can be calculated by making ⎾logn⏋ batch tests
- E(2^Y^) = Ɵ(k), i.e., E(2^Y^) is provably asymptotically proportional to k
- E(2^Y^) can be computed exactly given n and k
- Using linear regression we can find constants a and b such that p ≈ E((a2^Y^+b)/n)
- The variance of the estimator (a2^Y^+b)/n is O(p^2^). In contrast, the variance of the sample mean estimator is p(1−p)/⎾logn⏋. When p is small, our estimate is much more accurate than sampling individuals.

We will also define a random variable W such that

- W can be calculated by making m⎾logn⏋ batch tests, where m is a parameter
- W is the arithmetic mean of m independent copies of Y
- We will use W similarly to obtain a nearly unbiased estimator for p whose variance is O(p^2^/m).
- In practice this is better than using the arithmetic mean of m independent copies of 2^Y^.

Some identification algorithms based on batch testing are already known. We design two new highly parallel algorithms that are efficient for small p. The batch size for these algorithms is not fixed, but instead can be chosen optimally by making a calculation based on the estimated infection rate. We will describe how to choose an algorithm and its batch size given n and an estimate for k. One particularly favorable aspect of these algorithms is the fact that they use very few rounds of group testing which makes them easier to implement in practice than competing methods.

#### The Magic Trick Algorithm

To provide an intuitive example illustrating the principle of group testing methodologies we begin with a Magic Trick which is folklore in popular mathematics (Figure 1). We ask the reader to think of a number X between 0 and 31, e.g., X = 5. We now perform five binary tests on groups we carefully design. Each test returns 1 if the number X is in the group and 0 otherwise. In this case the result of the tests would be 00101 = 5, thereby identifying the hidden number. The tests and their results are listed below in Figure 1 for completeness.

**Figure 1:**
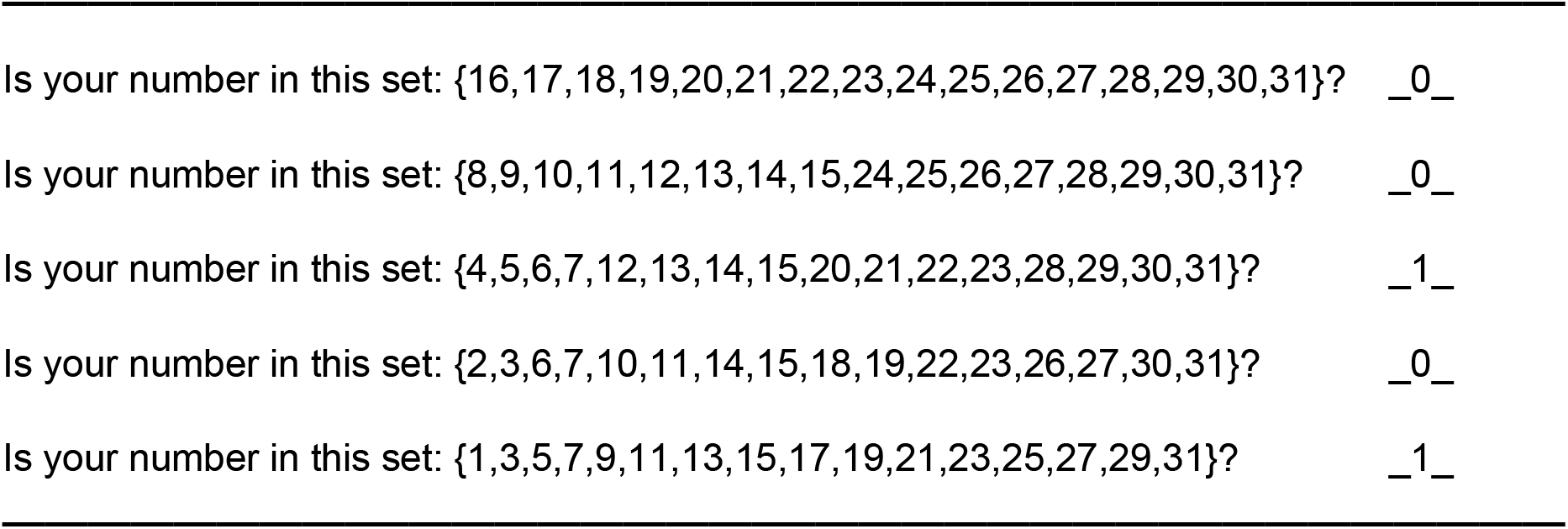
The Magic Trick: The answer is **00101 = 5**

The Magic Trick algorithm is described in full below.

Number the people 0 through n−1. Write those numbers in binary. Number the bits right to left starting from 0. (The i-th bit is in position 2^i^ in the binary representation. Alternatively, bit_i_(x) = (x div 2^i^) % 2.)

- Let m = ⎾logn⏋. All logarithms are base 2 unless otherwise specified.
- For i = 0 to m, let S_i_ = {x | bit_i_(x) = 1}
- For i = 0 to m, let b_i_ = [S_i_ tests positive]
- Let x be the number whose binary representation is b_m_b_m-1_…b_0_

Person number x is the one who is testing positive, as revealed by the magic trick above.

#### The Magic Trick Algorithm

Complexity Analysis: Each person sample is divided into ⎾logn⏋ samples. Exactly ⎾log⏋ tests are performed, and they can all be performed in parallel. The logistics of the process of producing the pools (batches) is not considered in this paper and can be performed by robotics or multi fluidic platforms. The size of the batches is n/2, which may potentially pose sensitivity and engineering challenges.

However, when k = n/2, information theory tells us that at least n − 0.5logn − 1 tests are required, so we cannot do significantly better than just testing every individual.

#### Rate of Infection Estimation by Approximate Counting Algorithms

We now describe approximate counting algorithms that use pools of samples to estimate accurate infection rates. We also provide sketches of the complexity analysis that provide bounds on the number of tests needed to estimate these rates. As alluded to before we are focusing on testing populations with a relatively low disease rate.

**Approximate Counting Algorithm (ACA1):** Number the samples randomly 1 through n. Choose independently subsets of size ⎾n/2⏋, ⎾n/4⏋, ⎾n/8⏋, ⎾n/16⏋, …, 1. Let Y = the number of subsets that test positive. Then EY ≈ log(k) where k is the number of infected individuals.

Complexity Analysis: Each person provides a single sample. ⎾log_2_n⏋ tests are performed, and they can all be performed in parallel.

The largest batch size is ⎾n/2⏋, which may pose some challenges as mentioned above. If k is not very small it is still possible to deal with the large batch size issue. For example, if the maximum allowed batch size is B then we could assume that all batches larger than B individuals would give the same test result as the largest batch. If all batches test negative, then we would estimate that the infection rate is less than 1/B. We can run ACA1 several times to produce a more accurate estimate W, which is discussed in the Results section.

### Section 2: Probabilistic Identification algorithms

In this section we will follow up on the rate estimation algorithms from the previous section and develop exact identification algorithms (both probabilistic and deterministic) of infected patients in a population with a low infection disease rate. We will present three algorithms, analyze their comparative performance and use each judiciously for the appropriate infection rate we estimated in the previous section. We note that Algorithm 0 is the natural approach that has been recently used in Nebraska with pool size s=5 without optimizing the choice of the pool size to reduce the number of tests. To be fair to this algorithm, sensitivity of testing in very large pools may be reduced without proper optimization and therefore we present Algorithm 0 (small pool size) below for completeness (Fig 2).

**Figure 2:**
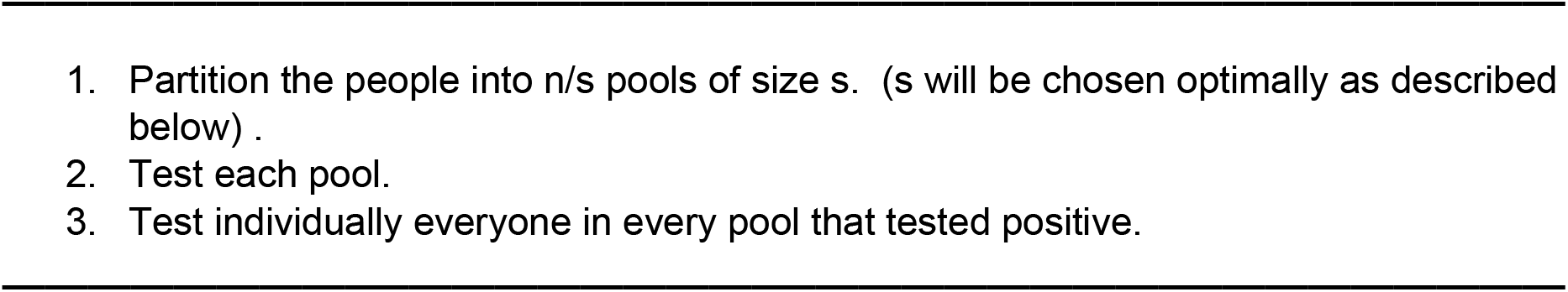
Probabilistic Identification Algorithm 0 (PIA0)

Analysis: The probability that a pool contains 0 positives is 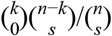. A pool that tests positive requires an additional s tests.Thus, on the average, Algorithm 0 performs 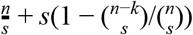 tests.

Our first new identification algorithm (PIA1) is given in Fig 3.

**Figure 3:**
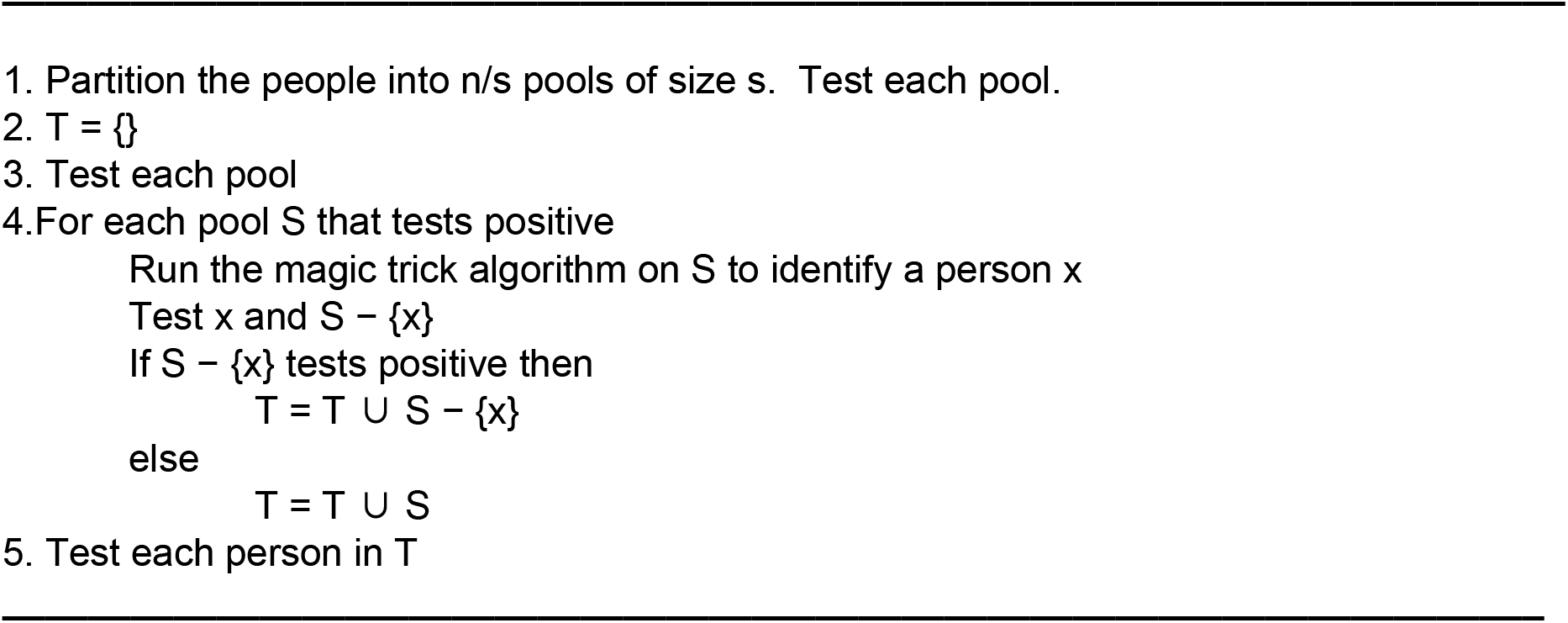
Probabilistic Identification Algorithm 1 (PIA1)

Analysis: The analysis of Algorithm PIA1 is given in the short summary below.

- The probability that a pool contains 0 positives is 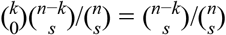.
- The probability that a pool contains exactly 1 positive is 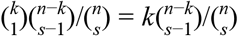.
- A pool that tests positive requires an additional ⎾log *s*⏋ + 2 tests.
- A pool with more than 1 positive requires an additional s tests.

Thus, on the average, Algorithm 1 performs 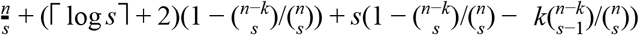 tests.

As an example consider this analysis with n = 10000, k = 100, and s=50. This is roughly a population of a small town with infection rate 0.01. We test 200 pools to start. On average 120.85 pools will contain 0 positives, 61.34 will contain exactly 1 positive, and 17.81 will contain more than 1 positive. We apply the magic trick algorithm on 79.15 pools of size 50, which takes 79.15log(50) = 474.9 tests. We check the results of the magic trick algorithm with 79.15 · 2 = 158.3 tests. Then we test the remaining 890.5 people. On average, the number of tests performed is 1723.7.

On average, 120.85*50 = 6042.5 people are classified in the 1st round, 61.34*50 = 3067 people are classified in the 3rd round, and the remaining 890.5 people are classified in the 4th round. On average, we obtain an individual’s test result in 1.88 rounds. We might test the remaining people recursively, but that is harder to analyze because the expected number of remaining tests is not linear in the number of remaining people. Recursion also increases the average and worst-case time to obtain an individual’s test result.

We would therefore use smaller pools than 50. For example, if we use pool size 25, we make only 1278.54 tests on average. In fact, pool size 24 is optimal. We calculated optimal pool size for several (n,k) pairs. Empirically, optimal pool size seems to depend primarily on k/n. When k/n = 0.01, optimal pool size is 24 or 25 in the examples we tried.

We now improve on the previous algorithms described in this section and present an improved Probabilistic Identification Algorithm 2 (PIA2) for a specific range of infection rate. The algorithm and its analysis are given below in Figure 4.

**Figure 4:**
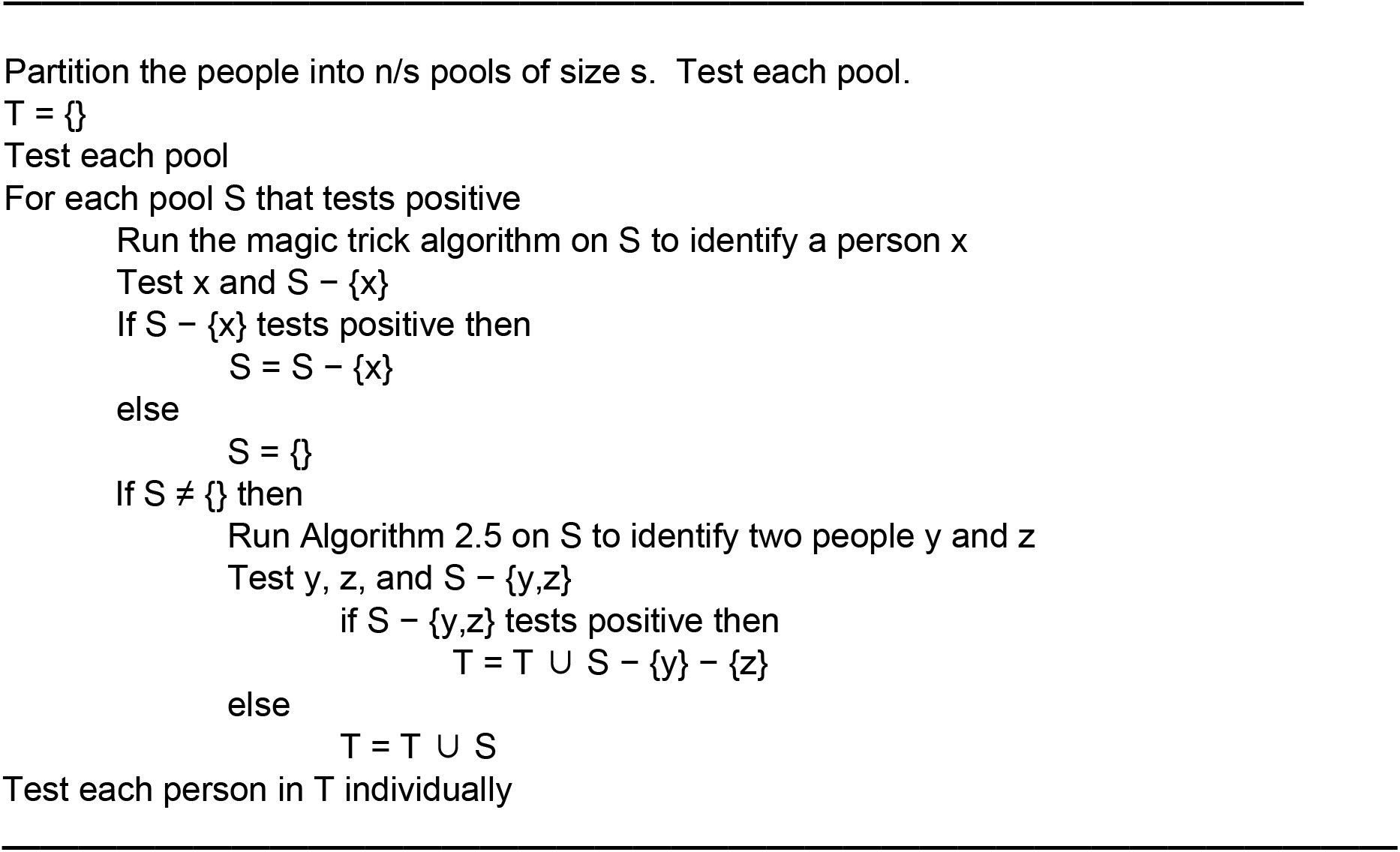
Probabilistic Identification Algorithm 2 (PIA2)

The analysis of the PIA2 algorithm is provided in the list below.

- The probability that a pool contains 0 positives is 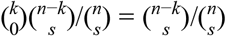.
- The probability that a pool contains exactly 1 positive is 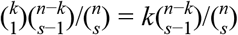.
- The probability that a pool contains exactly 2 positives is 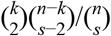.
- A pool that tests positive requires an additional ⎾log *s*⏋ + 2 tests.
- A pool with more than 1 positive requires an additional T(s)+3 tests, where T(s) is the number of tests performed by Deterministic Algorithm 2.5 when the population size is s.
- A pool with more than 2 positives requires an additional s tests.

To summarize, on the average, the number of tests performed by Algorithm 2 is:

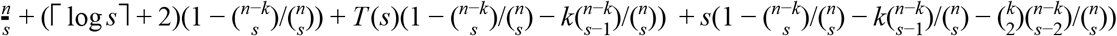

Given n and an upper bound on k, it is not difficult to implement a program that calculates T(s) as well as the expected number of tests made by Probabilistic Identification Algorithms 0, 1, and 2 for all s between 2 and n/k. The output of the program informs us which combination of algorithm and pool size makes the fewest tests on average. We provide a few illustrative examples below:

Example 1: consider: n = 10,000 and k ≤ 100. PIA2 with s=32 is the best choice. The expected number of tests is 1107.27. For comparison, Algorithm 0 performs 1956.59 tests on average with its optimal pool size.

Example 2: n = 100,000 and k ≤ 100. PIA2 with s=88 is the best choice. The expected number of tests is 2074.99. For comparison, PIA0 performs 6276.37 tests on average.

### Deterministic Identification algorithms for very small k

We now provide a deterministic identification algorithm applicable to populations with small infection rate (Fig 5).. A brief outline of the efficiency of the methods is provided in the list below.

- **Identification given k = 1:** ⎾logn⏋ tests in parallel (Magic Trick Algorithm)
- **Identification given k ≤ 1:** ⎾log(n+1)⏋ tests in parallel (left to the reader)
- **Identification given k ≤ 2:** 2.5logn − 1 tests in loglogn − 1 rounds (described below)

**Figure 5:**
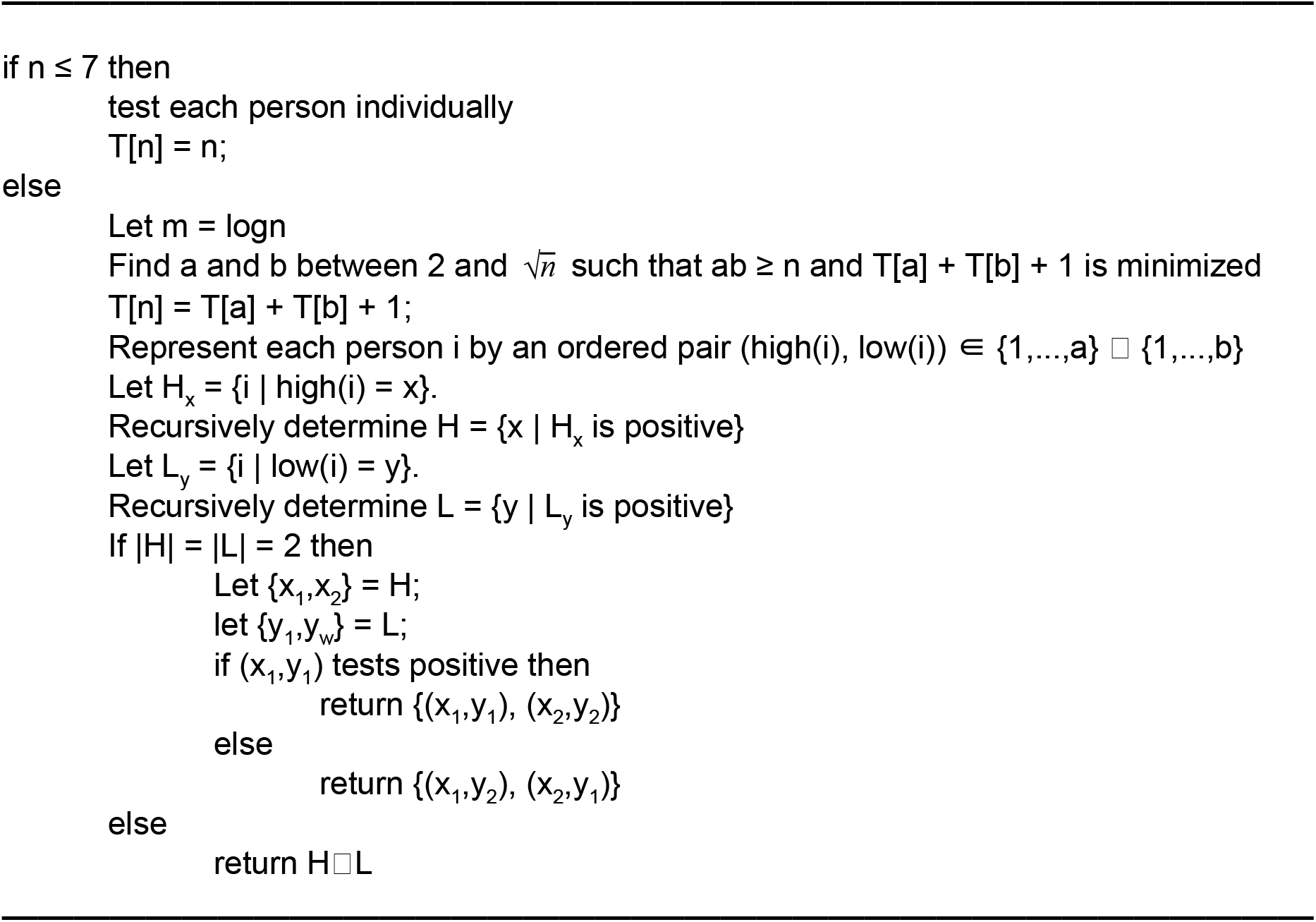
Deterministic Algorithm 2.5: Identification given k ≤ 2

Analysis: Let T(n) denote the number of tests made by DA2.5. Assume logn is a power of 2.

- 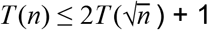
- T(4) = 4

By solving the recurrence we find T(n) ≤ 2.5logn − 1 when logn is a power of 2.

Example. What is T(36)?

- T(9) = T(3) + T(3) + 1 = 7
- T(12) = T(3) + T(4) + 1 = 8
- T(36) = T(3) + T(12) + 1 = T(4) + T(9) + 1 = 12

Note that T(36) < 2T(6) + 1.

Additional examples:

- T(16) = T(4) + T(4) + 1 = 9
- T(32) = T(2) + T(16) + 1 = T(3) + T(12) + 1 = T(4) + T(9) + 1 = 12

## Results

We first present a few empirical findings of approximate counting algorithms for infection rate estimation that generally support our ability to produce relatively accurate estimates using the methods provided in the previous section.

**Figure 6:**
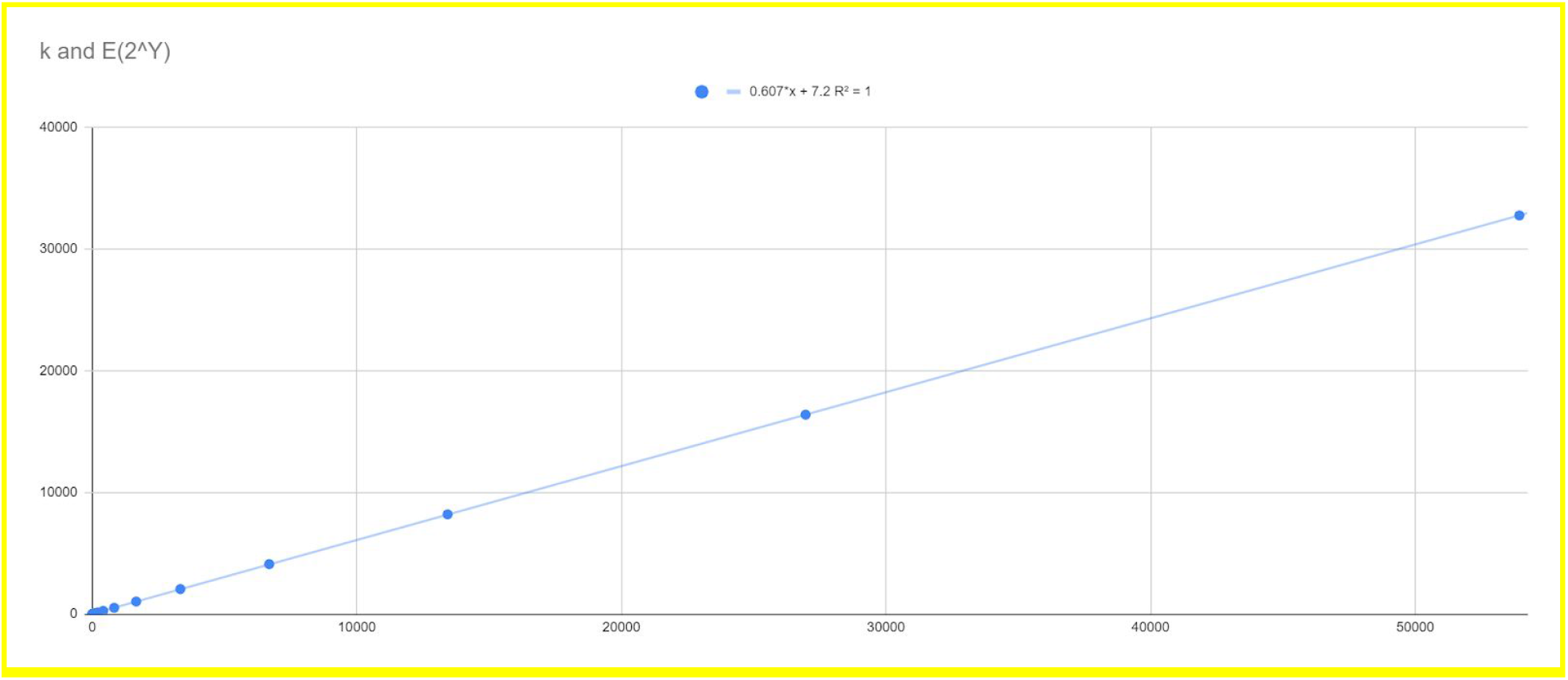
This graph displays the nearly linear relationship between k and E(2^Y^). In order to exploit this relationship in practice, we simply run a program that calculates E(2^Y^) for a known value of n and various values of k, and then perform a linear regression.

Figures 3-4 display the comparative accuracy of our pooling algorithms for estimating infection rates. In particular, Figure 4 demonstrates the reduction in variance obtained by running ACA1 multiple times.

In order to estimate the infection rate we run ACA1 m times. Let W be the average of those results. The number of infected individuals k is linear in 2^W^ We compute a linear regression to determine constants such that k ≈ a2^W^ + b. Then the infection rate, p, is approximately (a2^W^+b)/n. We refer to this algorithm as MACA1. (In practice it is better to perform linear interpolation using two adjacent data points. The linear formula, however, is needed in order to estimate variance.)

**Figure 7:**
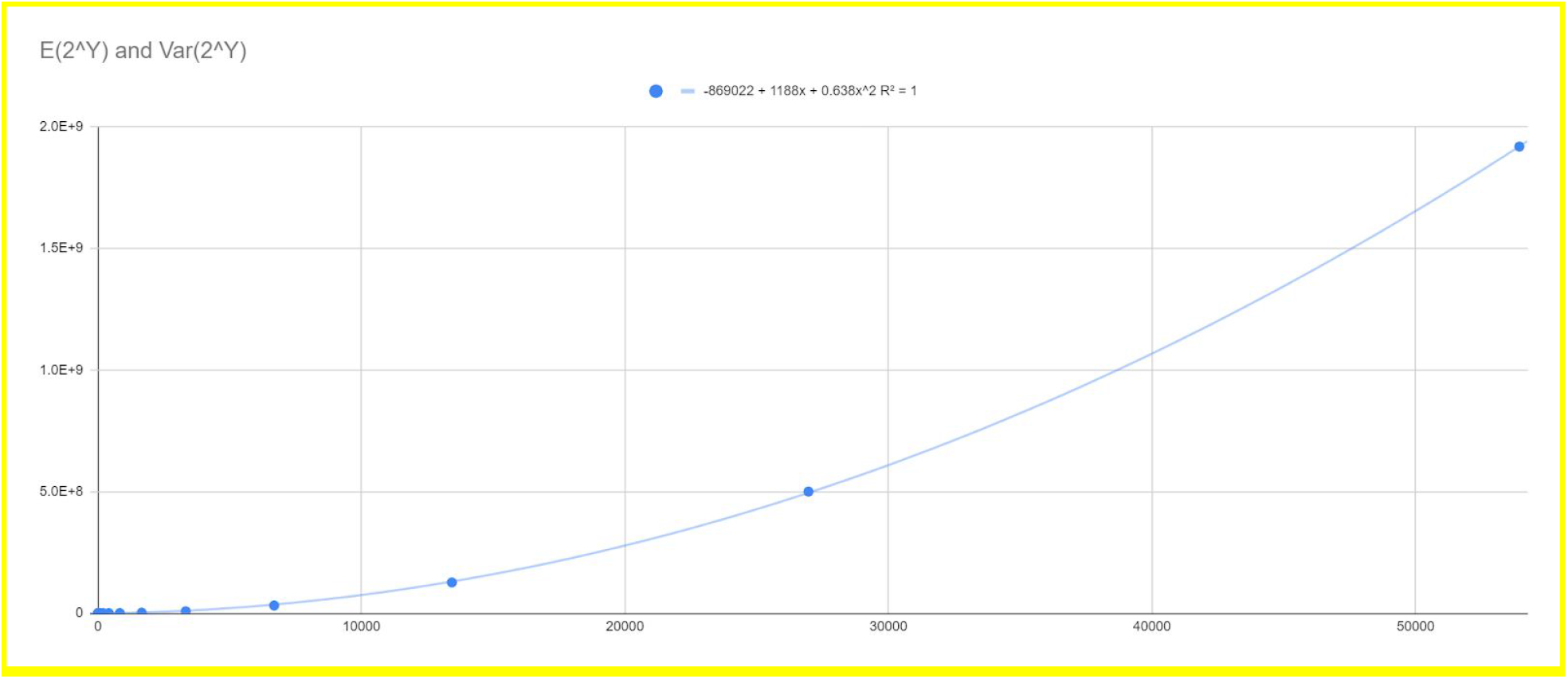
This graph shows that Var(2^^^Y) is approximately quadratic in E(2^^^Y) and the lead coefficient is less than one.

**Figure 8:**
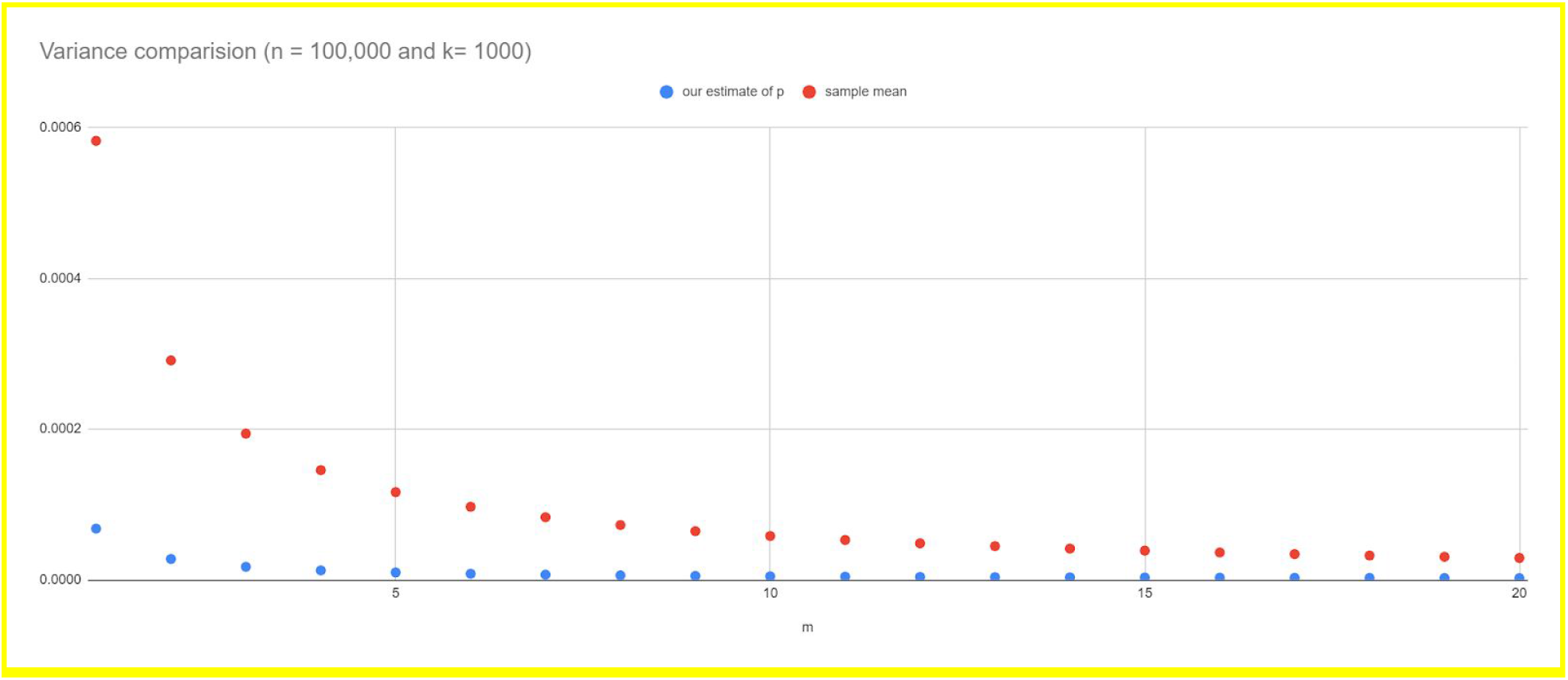
The blue dotted graph shows the variance of the MACA1 estimator. The red dotted graph shows the variance of the sample mean of m⎾logn⏋ tests.

### Empirical Findings of Probabilistic Identification Algorithms for Different Infection Rates

We now summarize the results of our probabilistic identification algorithms that we refer to as PIA#. We start by providing a sample of our empirical findings using a few selected examples in Table 1. The full graph is provided in Figure 9 for a relatively large population size.

**Table 1:** Testing efficiencies of the three algorithms: selected examples

| Infection Rate | Population Size | Best Algorithm |
| --- | --- | --- |
| $p = 0.061$ | $1000 \leq n \leq 10^6$ | PIA2 |
| $p = 0.062$ | $5500 \leq n \leq 10^6$ | PIA1 |
| $p = 0.069$ | $1000 \leq n \leq 10^6$ | PIA1 |
| $p = 0.07$ | $1200 \leq n \leq 10^6$ | PIA0 |
| $p = 0.3$ | $40 \leq n \leq 10^6$ | PIA0 |
| $p = 0.31$ | $300 \leq n \leq 10^6$ | Individual testing ($n$ tests) |

**Figure 9:**
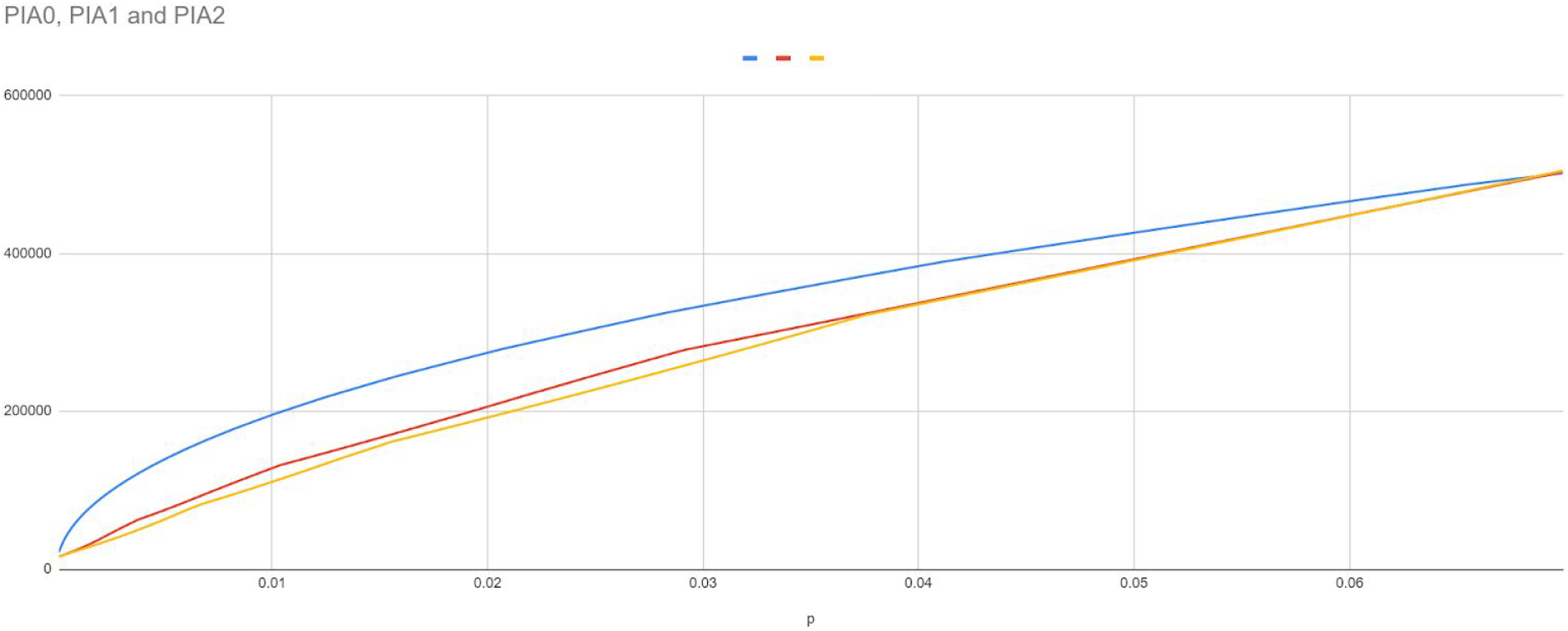
The expected number of tests performed by different algorithms for different infection rates p. PIA2 is graphed in yellow. PIA1 in red and PIA0 in blue. PIA2 generally outperforms the other algorithms when p ≤ 0.06. In this graph n = 1,000,000.

In Figure 9, we graphed the behavior (expected number of tests) of PIA0, PIA1, and PIA2 when n = 10^6^ and 0 < p < 0.07. We also observe that the optimal pool size seems to depend mostly on p. An information-theoretic lower bound on the number of tests required is 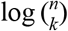. When p ≥ 0.005, we found that the number of tests performed is usually less then 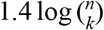.

## Discussion

The COVID-19 pandemic produced a very high toll worldwide and created technological, biomedical and clinical challenges. In this paper we described a cost effective application of approximate counting methods to estimate infection rates. These methods can help make crucial and rapid policy decisions with reduced investment, especially in early or late stages of the epidemic or in underserved communities, e.g., rural counties or third world countries. We also reviewed several identification algorithms and introduced new ones that provide good to modest improvements in the number of tests for different infection rates. In some cases our methods reduce the number of tests by two fold which is significant.

Estimating the infection rate without identification has many applications. These estimates can help policy makers determine appropriate guidelines for opening or closing the economy or implementing different types of social distancing procedures. The new method lends itself naturally to performing a very small number of tests on autopsy samples and is potentially useful in counting the number of individuals who were infected prior to death.

We also note that our proposed counting method can speculatively be extended to large scale vaccine trials. A large population can be vaccinated and another population can be used for placebo. We can use our cost efficient estimates to compare the rates in these two cohorts. In fact, we expect our variance estimates to be even better than we report above.

Our specific identification procedures of infected individuals improved on the number of tests that must be conducted saving in costs, especially when the infection rates are low and most tests are negative. Notably, we use a very small number of rounds as compared to the best competing algorithms, enabling quicker turnaround.

In this paper we primarily focused on using established methods such as PCR or isothermal amplification that have been shown to produce the most reliable diagnostics in the past. We have not considered multiplex-PCR as an alternative. There are newer diagnostic procedures based on CRISPR (CAS-13) under development. These methods vary in sensitivity, availability and cost. In theory, it is also possible to barcode every sample with a genomic tag, pool the tagged sequences into a very large multiplexed DNA sample and submit the sample for high throughput sequencing. Standard procedures would allow us to read the sequences and allocate any detected viral DNA to the appropriate individual using tags. Error correcting procedures can be deployed to produce an efficient library of tags. However, high throughput sequencing presents its own challenges and therefore we focused on pooling samples and testing using more traditional approaches. These established methods might also be easier to deploy in third world countries or rural communities that do not have access to high throughput solutions.

This work can be extended in multiple useful directions both mathematically and technologically. It is highly timely for a careful consideration and possible deployment given the expected flattening of the infection curve as we approach the summer of 2020 and the potential need to detect the disease in the fall of 2020.

## Data Availability

Self Contained

## Supplement: Formulas

The formulas in this section were used in

1. proving our theoretical results about expected value and variance of 2^Y^ and 2^W^
2. creating tables of E[2^Y^] for a given n (and all k) and E[2^W^] for given n and m (and all k)
3. creating tables of E[4^Y^] and E[4^W^]

The tables of E[2^Y^] and E[2^W^] make it possible to estimate k via linear interpolation. The tables are also used with linear regression to obtain our empirical findings about expectation and variance.

Formulas used with ACA1

- 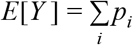
- where 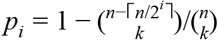
- 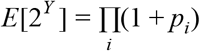
- 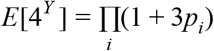
- *Var*(2*^Y^*) = *E*[4*^Y^*] − (*E*[2*^Y^*])^2^

Formulas used with MACA1:

- For any c > 0, 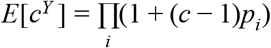
- *E*[2*^W^*] = (*E* [(2^1/^*^m^*)*^Y^*])*^m^*
- *E*[4*^W^*] = (*E* [(4^1/^*^m^*)*^Y^*])*^m^*
- *Var*(2*^W^*) = *E*[4*^W^*] − (*E*[2*^W^*])^2^

